# Catastrophic Health Expenditure Before and After of the Implementation of Health Sector Evolution Plan

**DOI:** 10.1101/2020.09.28.20202838

**Authors:** Razieh Ahmadi, Milad Shafiei, Hosein Ameri, Roohollah Askari, Hossein Fallahzadeh

## Abstract

**Objective:** One of the fundamental goals of health sector evolution plan (HSEP) in Iran is to improve household’s financial protection against catastrophic health expenditure (CHE). The aim of this study was to calculate the percentage of CHE after implementing the plan and compare that with CHE before the plan at the same households.

**Methods:** The data was collected through face-to-face interview during a single visit. The World Health Survey (WHS) questionnaire was completed by 400 households. The relationships between CHE and variables were examined by the Fisher exact tests, and the impacts of variables on CHE were assessed by logistic regression model.

**Results:** The exposure of the households to CHE increased from 8.2% in 2011 to 14.25% in 2020, and percentage of the impoverished households due to health expenditures in 2020 was more than that in 2011(4.3% vs. 7.5%). The economic status, dental services and inpatients services were the key factor determining CHE. The most important determinant affecting the exposure to CHE was dental service utilization in 2011(92.64) and 2020(122.68).

**Conclusion:** The results showed a negative incremental change for the households facing CHE in this period. The dental and inpatients services, as well as the presence of member ≥65 years and economic status were the key determining factors for CHE. The services need to be more widely covered by the basic health insurance and households having members ≥65 years and the poor households should be exempted from paying some of the healthcare expenditures for improving financial protection against CHE.

## Introduction

Increasing healthcare costs is a major concern in the most countries, especially in developing ones like Iran. The costs may impose a heavy burden on household and push many households into poverty. Hence, the world health organization (WHO) have a special emphasis on financial protection of households against healthcare costs and access to necessary health services without any financial constraints during receiving the service(1). The financial protection for the household occurs when household’s health out-of-pocket (OOP) payments are in proportion to its ability to pay, otherwise, some household will be exposed to catastrophic health expenditure (CHE)(2).

The rate of households’ exposure to CHE is one of the most commonly used indices to evaluate and control the financial protection status, and its use has recommended by the WHO and the World Bank(1, 3). The WHO defines CHE as a state when household’s health OOP is equal or exceed 40% of household’s income after paying for essential expenditure (4). The costs happen more in developing countries, where they rely more on OOP payments to finance health care. The share of health OOP payment for the total health expenditure in Iran was as high as 58%, and subsequently, the CHE rate was significantly high, as several regional studies reported the rate of households’ exposure to CHE between 8.3% to 22.2%(5). These features were not in line with the objectives intended in the Fifth Economic, Social, and Cultural Development Plan; that is, reducing the share of OOP payments to 30% in the total health expenditure and decreasing the percentage of households’ exposure to CHE to less than 1%. In addition to the problems mentioned, there were some other problems with Iranian health system, including low bed occupancy rate, informal payments, poor quality of care, inadequate response to increased demands, inequity access to healthcare services and insufficient insurance coverage. However, the Ministry of Health and Medical Education (MoHME) in 2014 has launched a series of reforms, called as the health sector evolution plan (HSEP). The plan followed three main objectives introduced by WHO, that is, improving health, enhancing responsiveness, and financial protection of households against healthcare costs. To achieve the goals, the packages, including many interventions, introduced by the MoHME. The majority of interventions focused on financial protection of households against healthcare, such as covering uninsured people by health insurance with no premium; decreasing coinsurance to 5% and 10% for, respectively, rural and urban residents to receive hospital services in public hospitals affiliated with the MoHME; eliminating informal payments through increasing medical tariffs; reducing the paid money amount by patients qualified for basic health insurance by 6% and 3% of the total hospitalization expenditures for urban and rural residents, respectively, and residents of cities with <20,000 population; providing all necessary medicines, consumables and services inside the hospitals; and reducing the number of unnecessary referrals to private centers. However, the plan funded by two main public sources, including 10% of incomes earned by the targeted subsidies initiative resources and 1% of the revenues obtained from value-added tax.

Nevertheless, this plan imposed heavy burden on the government and information about of the degree of success achieving the goals especially in financial protection is ambiguous. Several studies have evaluated the rate of CHE after implementation of HSEP(6-8), but only one study calculated the rate using a before and after approach(5). The study compared households’ exposure rate to CHE in the before (2008) and after (2015) of the implementation of HSEP using local data. Since the data obtained from 2015 was for lower than 1 years after implementing HSEP, it couldn’t show the real effects of HSEP on the financial protection of households. In line with lack of information, the goal of present study was to examine the rate of CHE before (2011) and after (2020) of the HSEP using local data.

## Method and Materials

In 2011, a survey was conducted in Yazd city, a city located in Central Iran with population close to one million. The survey had been carried out to determine the amount of households’ exposure to CHE and to identify factors associated with CHE among households. In 2020, the same households were re-examined and compared the changes. Neighboring households were selected instead of the households that could not be included in this study for any reason.

### Sampling and sample size

In 2011, a two-stage cluster sampling design had been used for selecting samples. In the first stage, 18 health centers, as 18 clusters, was selected based on geographical sampling frame which covered the entire population of city. The number of households in each of the clusters was calculated based on proportion of the population under coverage of each health center. In the second stage, the cluster head of each cluster was randomly selected based on information recorded for each people in each center.

In 2020, we followed the same households (or addresses) had been selected in 2011. Of the 340 addresses available from the 2011 survey, 43 households were missed for various reason such as changed street plates, building demolition, and households’ unwillingness to respond. Those replaced with neighboring households.

### Data collection

The World Health Survey (WHS) questionnaire developed by the WHO was used in both survey. The questionnaire included individual and household questions, and its validity and reliability have been confirmed in Iran(9). The data was collected through face-to-face interview in person’ home during a single visit. At the household level, a member ≥18 years old who was willing and able to answer the questions and had the most information about households’ general and healthcare expenditures was selected to complete the questionnaire. If, for any reason, the interviews could not be conducted with the selected household, the next nearest right-neighboring household was replaced. Data quality control was carried out through checking the collected data by telephoning 8% of the households. This study was approved by the SSU Ethics Committee, with the following identification: IR.SSU.SPH.REC.1398.095. The written informed consent was obtained from all participants prior to data collection.

### Statistical Analyses

Catastrophic health expenditures were calculated through the WHO approach. In parallel, health OOP payments equal or exceed 40% of capacity to pay determined as CHE(4). The relationships between the variables (economic status, health insurance status, having member with age higher than 65 years old, having member with age lower than 5 years old, having disabled member, dentistry services usage and inpatient & outpatient service usage) and CHE were examined by Fisher exact tests. In addition, the impacts of the variables on CHE were assessed by logistic regression model. Stata version 15 was used for data analyses.

## Results

The number of 43 households was replaced by neighboring households. Among households’ demographic and clinical variables, the percentage of member over 60 and disable number in households for 2020 were more than those for 2011. Mean all expenditures in 2020 were higher than those in 2011.

The exposure of the households to CHE increased from 8.2% in 2011 to 14.25% in 2020, and percentage of impoverished households due to health expenditures in 2020 was more compared to that in 2020(4.3% vs. 7.5%) (Table 2).

**Table 1.** Characteristics of both survey respondents

| **Variables** | **2011(396)** | **2020(400)** |
| --- | --- | --- |
| Household size | 4.11 | 3.24 |
| % households having member over 65 years | 3.01 | 3.51 |
| % households having member under 5 years | 28.43 | 23.18 |
| % households having disable number | 11.12 | 13.25 |
| % households having health insurance | 9.86 | 15.25 |
| Mean of household monthly total expenditure (IRR) | 14,324,000 | 31,804,510 |
| Mean of household monthly health expenditure (IRR) | 87,543,00 | 2,823,300 |
| Mean of household monthly dental services expenditure (IRR) | 5,854,100 | 1,597,400 |

**Table 2.** Percentage of households facing to CHE and impoverished households due to health expenditures

| **variables** | **2011(396)** | **2020(400)** |
| --- | --- | --- |
| % households exposed to CHE | 8.3 | 14.25 |
| % impoverished households due to health expenditures | 4.3 | 7.5 |

The Fisher exact tests showed that there were a significant relationship between facing CHE and variables of household size, economic status, dentistry usage, and inpatient service usage in 2020(P<0.1); and also showed that the relationships between facing CHE and variables of household size, member ≥65 years in household, economic status, dentistry usage, and inpatient service were significant (P<0.1) (Table 3).

**Table 3.** Relationship between the characteristics of households facing CHE and the variables in two surveys

| **Variables** | **Faced with CHE (2011)** |  | ***p*** | **Faced with CHE (20220)** |  | ***p*** |
| --- | --- | --- | --- | --- | --- | --- |
|  | Yes (%) | No (%) |  | Yes (%) | No (%) |  |
| **Household size** |  |  | 0.04 |  |  | 0.06 |
| ≤2 | 4(4.3%) | 89(85.7%) |  | 9(8.8%) | 93(91.2%) |  |
| 3-6 | 28(9.2%) | 275(81.8%) |  | 48(16.1%) | 250(83.9%) |  |
| **Member ≥65 years in Household** |  |  | 0.09 |  |  | 0.72 |
| Have | 8(36.3%) | 14(73.7%) |  | 2(14.2%) | 12(85.8%) |  |
| Not have | 24(6.4%) | 350(93.6%) |  | 55(14.2%) | 331(85.8%) |  |
| **Member ≤ 5 years in Household** |  |  | 0.44 |  |  | 038 |
| Have | 7(8.5%) | 75(91.5%) |  | 11(11.7%) | 84(88.3%) |  |
| Not have | 25(7.9%) | 289(82.1%) |  | 46(15.1%) | 258(84.9%) |  |
| **Disabled member** |  |  | 0.32 |  |  | 0.49 |
| Have | 6(13.6) | 38(86.4) |  | 8(14.04) | 45 (13.12) |  |
| Not have | 26(7.4) | 326(92.6) |  | 49(85.96) | 298(86.88) |  |
| **Economic status** |  |  | 0.001 |  |  | 0.02 |
| Quintile 1 (poorest) | 12(13.9%) | 74(86.1%) |  | 21(21.4%) | 77(78.6%) |  |
| Quintile 2 | 8(5.7%) | 131(84.3%) |  | 24(16.5%) | 121(83.5%) |  |
| Quintile 3 | 6(12.2%) | 43(87.8%) |  | 2(7.7%) | 24(92.3%) |  |
| Quintile 4 | 4(5.9%) | 63(94.1%) |  | 7(9.3%) | 68(90.7%) |  |
| Quintile 5 (richest) | 2(3.6%) | 53(96.4%) |  | 3(5.3%) | 53(94.7%) |  |
| **Health insurance status** |  |  | 0.11 |  |  | 0.36 |
| Have | 12 (20.7%) | 46(79.3%) |  | 10 (17.5%) | 51(14.9%) |  |
| Not have | 20 (7.3%) | 318(92.7%) |  | 47 (82.5%) | 292 (85.1%) |  |
| **Inpatient service usage** |  |  | 0.09 |  |  | 0.04 |
| Have | 9(30%) | 21(97%) |  | 7(26.9%) | 19(73.1%) |  |
| Not have | 23(6.3%) | 343(93.7%) |  | 50(13.3%) | 324(86.7%) |  |
| **Outpatient service usage** |  |  | 0.13 |  |  | 0.84 |
| Have | 7(7.8%) | 82(92.2%) |  | 15(14.8%) | 86(84.2%) |  |
| Not have | 25(8.1%) | 282(91.8%) |  | 42(14%) | 257(86%) |  |
| **Dentistry usage** |  |  | 0.001 |  |  | 0.001 |
| Have | 23(44.2%) | 29(55.8%) |  | 42(56.7%) | 32(43.3%) |  |
| Not have | 9(2.6%) | 335(97.4%) |  | 15(4.6%) | 311(95.4%) |  |

Results from the logistic regression model, where households facing CHE was as a dependent variable and the variables of member ≥65 years in household, member ≤5 years in household, having health insurance, economic status, inpatient service usage, outpatient service usage, and dentistry usage were as independent variables, showed that only effect of having member ≥65 years in household changed to significant in 20120 (Table 4). The chance of households’ exposure to CHE in the household with members over 65 years was OR=1.49 (0.05, 4.10) in 2020. The chances of facing CHE for the rich households (0.01325) was lower than other households ranged between 0.97843 and 0.15468 in 2011; and the rich households also had the lower chance of exposure to CHE compared to other households ranged between 0.26518 and 0.06468 in 2020. The chances of exposure to CHE in the households taking inpatient services, outpatient services, and dental care services were 6.47, 1.42, and 92.68 in 2011, respectively, and the households receiving these services had the chance of facing CHE of 8.19, 2.39, and 122.68, respectively, in 2020.

**Table 4.** Determinants of catastrophic health expenditure in the logistic regression model

|  | **OR (2011)** | **95% CI** | **OR (2020)** | **95% CI** |
| --- | --- | --- | --- | --- |
| **Member ≥65 years in Household** | 1.18943 | [0.01889 3.65062] | 1.49184* | [0.058911 4.10632] |
| **Member ≤ 5 years in Household** | 1.07244 | [0.93254 5.45674] | 1.54114 | [0.588208 4.03789] |
| **Having health insurance** | 1.93122 | [0.34101 4.79215] | 1.51516 | [0.524012 4.38102] |
| **Economic status** |  |  |  |  |
| Quintile 2 | 0.97843 | [0.22415 1.89357] | 0.26518* | [0.09419 0.74658] |
| Quintile 3 | 0.32651* | [0.13252 0.98214] | 0.02881* | [0.00242 0.34309] |
| Quintile 4 | 0.15468* | [0.01606 0.26041] | 0.06468* | [0.01606 0.26041] |
| Quintile 5 (richest) | 0.01325* | [0.00478 0.09872] | 0.00811* | [0.00124 0.05268] |
| **Inpatient service usage** | 6.47924* | [2.17524 14.9682] | 8.19720* | [2.25561 29.7890] |
| **Outpatient service usage** | 1.42831 | [0.35452 5.31455] | 2.39739 | [0.89031 6.45562] |
| **Dentistry usage** | 92.6457* | [23.7854 227.112] | 122.685* | [40.7671 369.213] |
| **constant** | 0.15743* | [0.08845 1.24685] | 0.08190* | [0.03706 0.18099] |

\*P < 0.05

## Discussion

The present study assessed the rate of the households’ exposure to CHE and the effects of socio-demographic and clinical factors on CHE before and after of the HSEP. The results of our study provided important insights of the CHE based on households’ characteristics that can be more appropriate for policy makers.

Mean CHE rate for households in 2011 and 2020 were as high as 8.3 and 14.25, respectively. The rates were higher than results (3.91%) obtained from a systematic review and meta-analysis carried out on the pooled date 1995 to 2015 in Iran. The findings also fell in the high range of 8.3% to 22.2% of the CHE in several regional studies conducted using the WHO questionnaire at level of Iranian households between 2009 to 2014. All findings showed that the rate of households facing CHE is high and is not in line with the Fifth Economic, Social, and Cultural Development Plan aiming to reduce the proportion of households facing CHE to less than 1%.

We observed that the proportion of households facing CHE, despite there is the major reform in this period, is increased of 8.3 to 14.25. The only other regional study conducted similar to our study in 17the district of Tehran, capital of Iran, showed that the percentage of households’ exposure to CHE increased from 11.8 in 2008 to 29.9 in 2015. While it was expected to observe significant reduction in CHE according to the data of the study collected in first year after implementation of the HSEP. In addition to increase in percentage of households’ exposure to CHE, the percentage of increase of CHE (153%) was higher than that (71%) in this study(5). One possible reason for this finding can be explained by different characteristics of the population of the district 17 of Tehran compared with the Yazd population. The districts’ population was associated with low socioeconomic status, high rate immigrants and work of many residents in hard physical occupations(5, 9). The increase in percentage of CHE after implementing the HSEP observed also in study carried out on national survey data by Yazdi-Feyzabadi et al.(7). They observed the exposure rate to CHE was increased from 1.99 in 2011 to 3.46 in 2017 in total urban and rural populations. The increase rate of CHE in the study (73%) was almost similar to that of 71% in our study(7).

One reason for the high rate of CHE after implementing HSEP in the province of Yazd and the above-mentioned studies could be attributed to the increase in public health tariffs aiming to make the tariffs closer to the actual final prices. Some empirical evidence showed that the proportion of OOP payments was reduced, but the final OOP payments did not decrease due to the sharp increase in public health tariffs(5, 10). Whilst according to the WHO report (2010), the negligible CHE occurs when that the OOP drop by 15% in a country(11). Yardim et al. observed that the rate of CHE after implementing the Health Transformation Programme in Turkey decreased to 0.6. They presented that decreasing OOP payment by 19.3% after the plan was one of the most important reason for diminishing CHE (12). A major part of the high rate of households’ exposure to CHE is related to the high annual inflation rate in the Iran’s economy, especially in recent years. The average annual inflation rate between 2015 and 2019(20.58) was higher than that of 2010 to 2014(18.64). In addition, the percentage increase of the annual inflation rate from the beginning of the plan to until the end of 2019 was 246% (13). This inflation has led to a sharp rise in the prices of goods and services and, consequently, an increase in health costs. As when comparing the mean expenditures of the household health services in 2020 with those in 2008, the mean health expenditure was almost 3 times higher than that in 2011. Another reason of the difference in CHE rate between before and after of the plan can be due to the increasing application of the new diagnostic and medical technologies in Iranian health system in recent years. For example, the use of MRI in diagnosis services was not so common in 2011 while in recent years, it is highly used.

The findings showed that only impact of having member ≥65 years in household on CHE changed to significant in this period. A systematic review and meta-analysis reported having a member aged 60–65 years or older in the household in 12 of 40 studies increased levels of CHE in Iran. These members usually are not employed in practice and do not earn income as well as, on the other hand, the age-related comorbidities are more in older adults. Therefore, presence of such members makes households spend a greater part of their capacity to pay on healthcare expenditures. The positive relationship between presence of the member ≥65 years in household and more likelihood of facing CHE was observed in some other countries, such as Turkey(12).

With regards to the features of insurance, it is expected to health insurance there was a protect effect for household against CHE. Nevertheless, this effect was not observed in this study. Some evidence of regional studies at level households in Iran support this finding. One reason for failure of health insurance may be because the insurance increases utilization of health care, and subsequently, the risk of CHE. This hypothesis that insurance coverage increases health care usage is supported by some studies (2, 14, 15). These findings suggest that more resources need to be allocated on improving the pattern of patient health care usage, managing provider for reducing supplier-induced demand or unnecessary care, and providing health care insurance.

The household economic status was a key factor determining CHE in both surveys, as households with better economic status were associated with lower chance of exposure to CHE. This reverse relationship between economic status and facing CHE were founded in the majority of studies in Iran (8, 9, 16) and other countries (12, 17, 18). Hospitalization is another one of the significant predictors of CHE in both surveys. The households receiving inpatient services in 2020 had more chance of facing CHE (OR=8.19) compared to that in 2011(OR=6.47), while it is expected that the rate of exposure to CHE after implementing the plan to be lower in comparison with that in 2011. This finding was consistent with that of the other study that was conducted before and after of the plan in Iran(5). One possible reason can be due to more use of the private sectors that their services are expensive and were not covered by the plan.

We found dental care usage is the most important determinant of CHE in both surveys. Dentistry services are among the more expensive health care services in Iran and are not usually covered by basic health insurance benefit packages. Most people pay directly out-of-pocket for receiving the services. This finding support by other studies were conducted in Iran(5, 9) and other countries, such as Brazil (17) and Korea(18).

The present study has some limitations that should be noted. First, the participants were restricted to the Yazd province, center of Iran, who they may not be perfectly representative of other Iranian populations. Thereafter, our results should be applied with caution. Second, recall period of expenditures and usage of services was shorted to minimize the recall bias.

## Conclusion

This study showed that proportion of households facing CHE, despite implementation of the reforms in this period, has increased from 8.4 in 2011 to 14.6 in 2020. Similar to other studies comparing the rate of CHE before and after of the plan we also found that the objective emphasized in the Fifth Economic, Social, and Cultural Development Plan (ie, reducing the proportion of households facing CHE to less than 1%) is not yet achieved.

The findings of this study and other studies in Iran indicated that dental services and inpatients were the key determining CHE and those need to be more widely covered by the basic health insurance. The results also showed the presence of member over 65 years old in household and households with low economic status led to significant increase in the chances of facing CHE. In this regard, households having member ≥65 years and the poor households should be exempted from paying some of the healthcare expenditures for improving financial protection against CHE.

## Data Availability

Data is available from the corresponding author on reasonable request

## Acknowledgements

The authors hereby bestow much gratitude to the households for their valuable collaboration and participation in the present study.

## Author contributions

HA conceptualized the study and the design of study. HA, RA and HF participated in data analysis and data interpretation. HA, MS, and RA wrote the original draft of the manuscript. All authors read and approved the final manuscript.

## Compliance with Ethical Standards Ethical Information

This report is part of a MSc project that was approved by the committee of Shahid Sadoughi University of Medical Sciences (approval number: IR.SSU.SPH.REC.1398.095).

## Declaration of Interest

The authors have no other relevant affiliations or financial involvement with any organization or entity with a financial interest in or financial conflict with the subject matter or materials discussed in the manuscript apart from those disclosed.

## Informed consent

Written informed consent was obtained from all participants included in the study.

## Conflict of interest

The authors declare that they have no conflict of interest.

## Funding

Finding for this project was provided from the Shahid Sadoughi University of Medical Sciences (registration number: 6837).

## Notes

### Competing Interest Statement

The authors have declared no competing interest.

### Clinical Trial

Not clinical trial

